# COVID-19 epidemic in Sri Lanka: A mathematical and computational modelling approach to control

**DOI:** 10.1101/2020.04.21.20073734

**Authors:** WPTM Wickramaarachchi, SSN Perera, S Jayasignhe

## Abstract

The ongoing COVID19 outbreak originated in the city of Wuhan, China has caused a significant damage to the world population and the global economy. It has claimed more than 50,000 lives worldwide and more than one million of people have been infected as of 04^th^ April 2020.

In Sri Lanka, the first case of COVI19 was reported late January 2020 was a Chinese national and the first local case was identified in the second week of March. Since then, the government of Sri Lanka introduced various sequential measures to improve social distancing such as closure of schools and education institutes, introducing work from home model to reduce the public gathering, introducing travel bans to international arrivals and more drastically, imposed island wide curfew expecting to minimize the burden of the disease to the Sri Lankan health system and the entire community. Currently, there are 159 cases with five fatalities and also reported that 24 patients are recovered and discharged from hospitals.

In this study, we use the SEIR conceptual model and its modified version by decomposing infected patients into two classes; patients who show mild symptoms and patients who tend to face severe respiratory problems and are required to treat in intensive care units. We numerically simulate the models for about five months period considering three critical parameters of COVID transmission mainly in the Sri Lankan context; efficacy of control measures, rate of overseas imported cases and time to introduce social distancing measures by the respective authorities.

## Introduction

Sri Lanka periodically faces epidemics of infections that cause morbidity and mortality. There is epidemiological data on specific diseases such as leptospirosis and dengue. Though clinicians observe periodic epidemics of influenza and other respiratory illnesses there is little scarce information on their spread, case fatality rates of numbers. This is partly because respiratory illnesses are quite common and regular diagnostic tests are rarely available in the state sector hospitals. Nevertheless, Sri Lanka seems to have escaped major viral epidemics respiratory illnesses such as Severe Acute Respiratory Syndrome (SARS) and Middle East Respiratory Syndrome (MERS) until, COVID-19 was detected in Sri Lankan tour guide on the 10^th^ of March 2020.

Though we have escaped SARS and MERS are coronaviruses that have caused two large-scale pandemics in the past two decades [1, 2]. Since the SARS epidemic outbreak of SARS, 18 years ago, a large number of similar viruses have been discovered in bats, its natural reservoir host, bats [3]. Around 12^th^ December 2019, a new coronavirus SARS-CoV -2 started an epidemic of acute respiratory syndrome in humans in Wuhan, China. Genetic sequencing has confirmed that this arose from the bat coronavirus [4].

The virus spreads from human-to-human via droplets or through contaminated surfaces which in turn enter the nasal mucosa, oral cavity or mucosa eyes through touch. This is similar to the spread influenza (e.g. H1N1) and SARS. However, COVID-19 potential of transmission is much higher [5]. As a result, the estimated doubling times ranges from 6.4 to 7.4 days [6].

Disease is characterized by cough, fever and sore throat and may result in virus-induced pneumonia and progressive respiratory failure owing to alveolar damage caused by the virus. As a result, the mortality rates from the illness also relatively high. The disease is also characterized by a mean incubation period was 5.2 days (95% confidence interval [CI], 4.1 to 7.0) [7].

The proportion of asymptomatic cases reached 30.8% (confidence interval 7.7% to 53.8%) [8]. It is found that the virus is shed throughout the illness. As a result, the virus is more infective for a longer duration than other illnesses such as. Milder cases cleared the virus by 10 days while 90% of severe cases continued to shed the virus in the nasal secretions beyond 10 days from onset of illness [9, 10, 11].

Briefing the COVID19 situation in Sri Lanka, the index case was a Chinese tourist who was detected with the illness on 27^th^ of January and admitted to the National Infection Disease Hospital. The first Sri Lankan patient known to be a tourist guide who had not traveled overseas was detected to have the illness on the 11^th^ of March. Having understood the potential spread of the disease over the entire population is Sri Lanka, the government of Sri Lanka took immediate sequential measures such as island wide school closure, Travel ban from selected countries (South Korea, Italy and Iran), declaration of special holidays to limit public gathering, Shutting down the Colombo International Airport for all arrivals to the island and finally decided to impose island wide curfew. As of 04th April, there are 159 confirmed cases of COVID19, 5 deaths and 24 recoveries [12].

Since Sri Lanka have been a developing country, the facilities to treat COVID19 patients are found to be very limited. Therefore, it is very critical to understand how sequential control measures should be introduced and in which capacity and efficacy level so that the epidemic curve can be flatten and it is possible to avoid the health system of the country overwhelmed [12, 13].

Mathematical modelling in epidemic diseases has a long history and many researchers adopted various types of models to simulate and predict the outbreaks [13]. In this study, we aim to develop a SEIR (Susceptible, Exposed, Infected, and Recovered) mathematical model and we also introduce an intermediate class of patients who are likely to become critically ill from the disease seek intensive care treatments [17]. Scenario based control measures are developed and they are introduced to the mathematical model. The numerical simulations are carried out to show how these measures are effective and also to predict the dynamic for a shorter period time so that the health system can prepare accordingly.

## Material and Methods

We apply ‘Susceptible-Exposed-Infected-Recovered’ (SEIR) framework to model the dynamic of COVID19 outbreak in Sri Lanka. We introduce two models basically that is the first model is a basic SEIR model using the traditional approach without considering the demography of patients. However, in the second model, we introduce the new class of cases includes the patients who may receive the intensive medical care due to the severity of the infection they undergo.

## Mathematical Models

### Model 1: SEIR model considering the homogeneity of patients

In this mathematical model, we assumed all the clinically tested positive patients for COVID-19 virus are homogeneous with no impact of age, gender and history of chronic diseases on the disease progression. Natural birth and death process was considered to be negligible and those who recovered to have developed complete immunity against the virus. In this simple model, the susceptible individual may become exposed to the novel corona virus at a rate of *β*. After passing the time to show the symptoms of COVID19 or time to identify clinically as positive to the virus σ , this exposed individual is designated as infected. As a result of treatments or due to the immunity, the patient can be recovered at a rate of *γ*, however, the patients whose condition is critical can end up losing their lives at a rate of *μ*. In the context of Sri Lanka, the island got the virus mainly from the exposed people who arrived from the overseas countries. Thus, we let λ be this rate. This dynamic can now be represents as follow in a schematic diagram (Figure 1).

**Figure 1:**
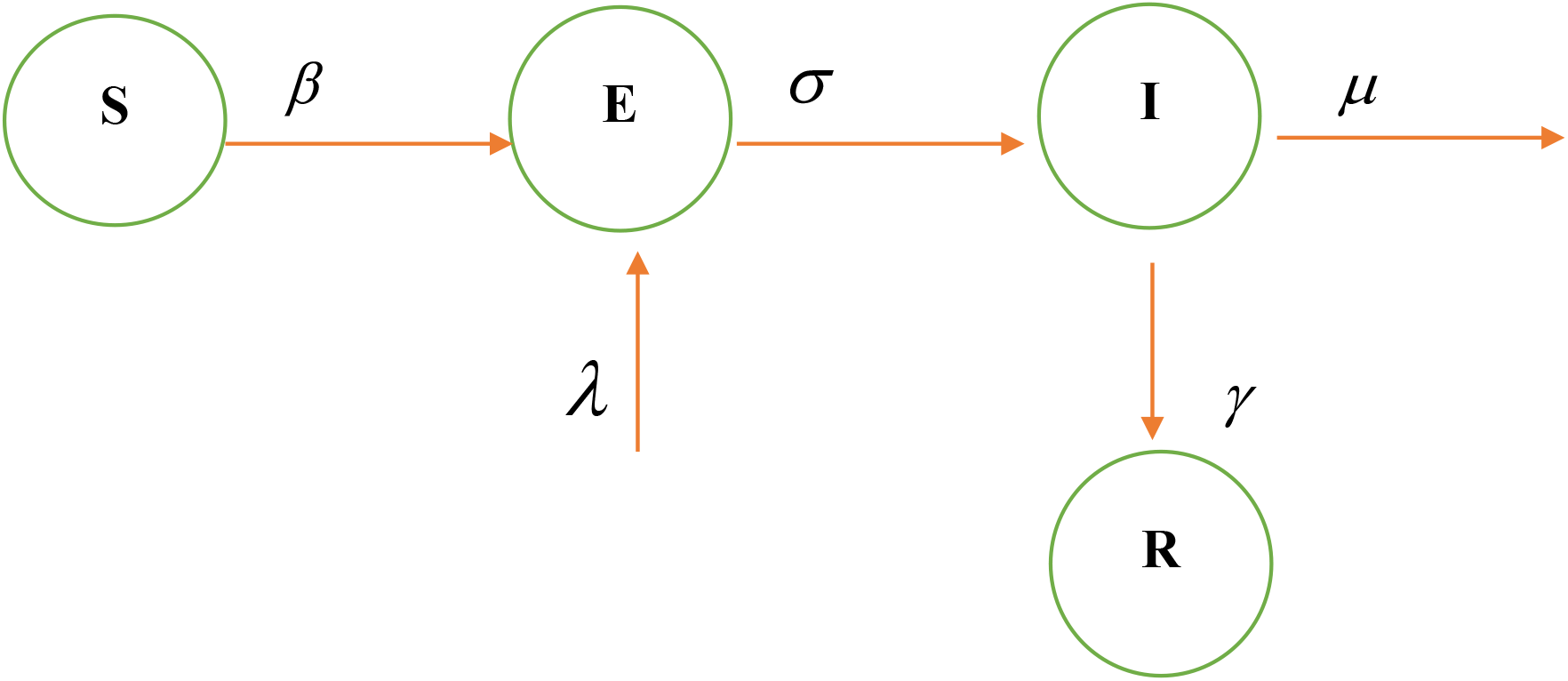
Schematic of model 1

The SIER (susceptible, Exposed, Infected and Recovered) compartment model can be described as follows;

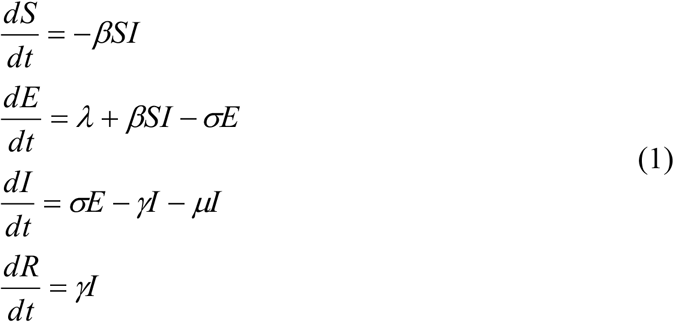

with non-negative initial conditions (*S*_0_, *E*_0_, *I* _0_, *R*_0_).

#### Deriving the Basic Reproduction Number (*R*_*0*_)

The Disease Free Equilibrium (DFE) is basically given by (*S*_0_, 0,0,0). According to the model in system (1), there are two infected classes of humans (*E* and *I*). We use the next generation matrix method to find the basic reproduction number (*R*_0_) described in [14]. We derive the gains to *E* and *I* classes respectively as *βSI* and *σE*, similarly the losses from *E* and *I* classes as *σE* and (*γ* + *μ*)*I* respectively. Now we define the two matrices *F* and *V* such that

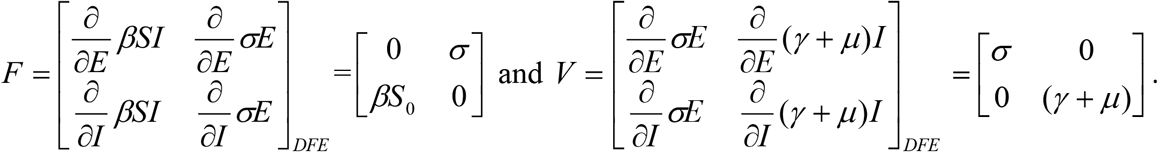

Now *FV* ^−1^ is the next generation matrix and *R*_0_ = *ρ*(*FV* ^−1^) where *ρ* is the spectral radius. Thus for the system (1), the basic reproductive number is derived as 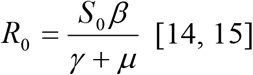 [14, 15].

#### Simulation of Model 1

The numerical simulation of the conceptual models for COVID19 transmission is carries out in MATLAB and the outcomes are shown in Figure 2. It should be noted that a hypothetical value for the transmission probability *β* is used for the simulation and we no control measures are included for the simulation. According to the simulation of model 1, the nearly 55 days after the first local patient is found, the curve attains to its peak.

**Figure 2:**
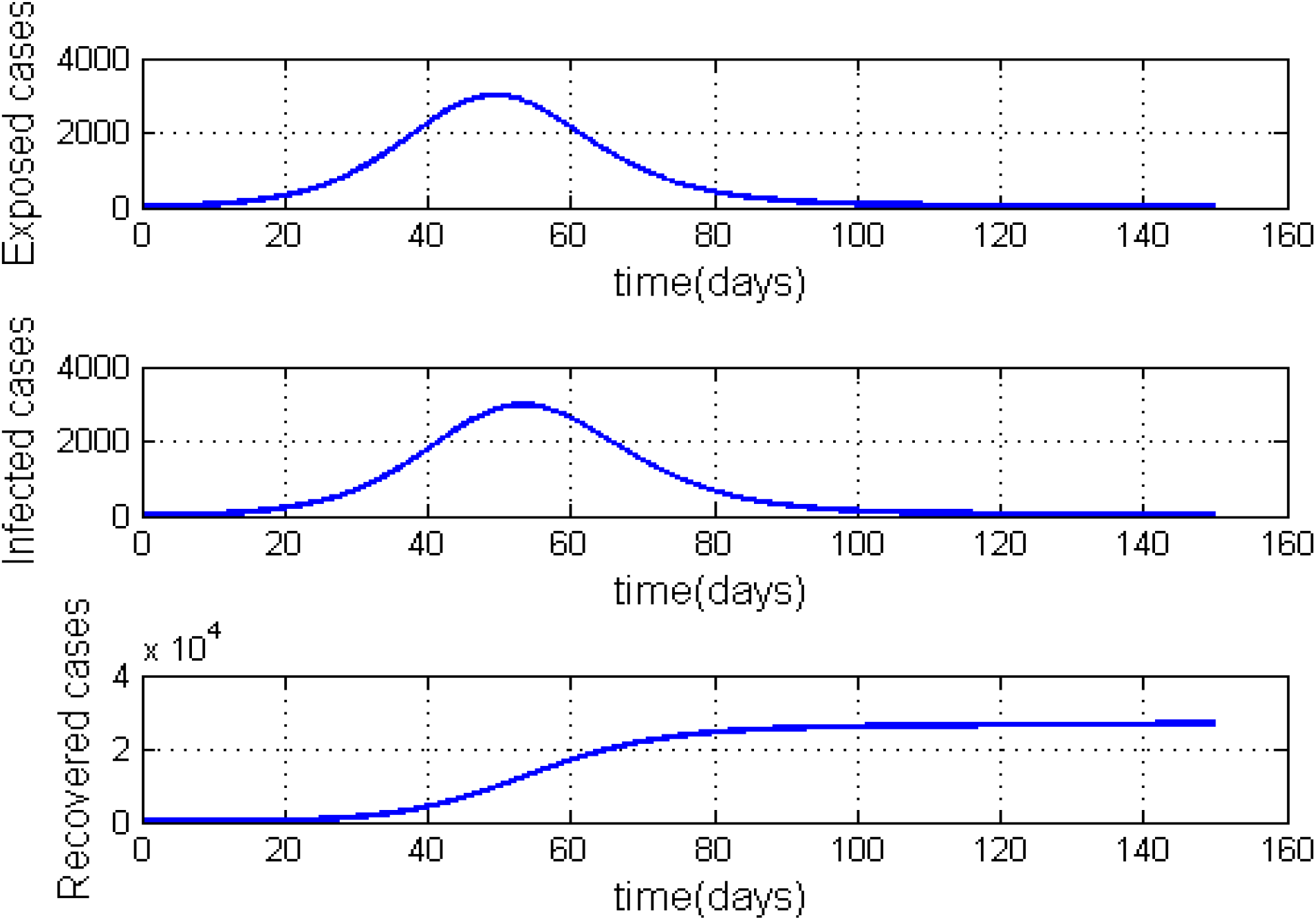
Simulation of model 1 without any control measures. Parameter values used for the simulation are *β* = 0.7, *γ* = 0.24, *μ* = 0.001, *σ* = 1/ 4 and *λ* = 0.000205[13].

### Model 2: *SEI*_*m*_*I*_*c*_*R* model considering the heterogeneity of patients (including the class of patients who will require ICU treatments)

In this mathematical model, we have assumed the clinically tested positive patients for COVID19 are not homogeneous and took into consideration their age, gender and history of chronic diseases that worsen severity and lead to ICU treatment [17]. The new group of patients *I*_*c*_ is added to the model representing the severe cases of COVID19. These severe patients are absorbed by the available ICU beds until it reaches capacity.

According to this model, the recovery can happen in two ways. First, the patients in *I*_*m*_ class may show mild symptoms of COVID-19 and eventually all of them recover fully. Second, the condition may become severe based on the patients age, gender, life style (smoking or alcohol addicted) presence of chronic diseases. However, they recover due to the ICU treatment they receive while a small proportion die. While all other parameters are the same, the new parameters are introduced to the model such that *δ* is the rate at which a patient’s level becomes critical. This should be in a measurable functional form with respect to several demographic variables. Further, *γ*_1_ is the recovery rate of patients who show mild symptoms while *γ*_2_ is the recovery rate of severely ill patients who are treated in ICUs where *γ*_1_ > *γ*_2_. In this model, the parameter *u* ∈ (0,1) is the control parameter and it depends on multiple control and social distancing variables. It is further assumed that the patients in ICU are isolated from the public and they are not responsible for the community transmission [13, 17]. The dynamic can now be represented according to the following schematics diagram (Figure 3).

**Figure 3:**
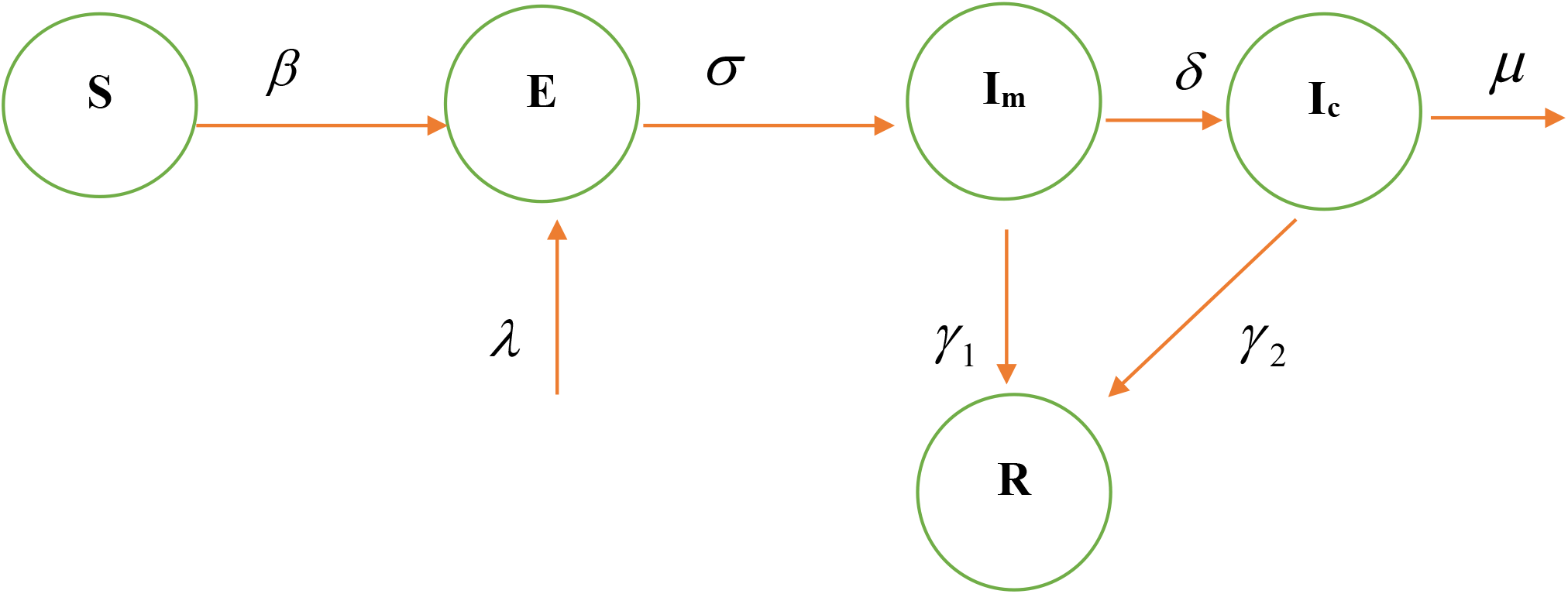
Schematic of model 2

The new model can now be stated as follows:

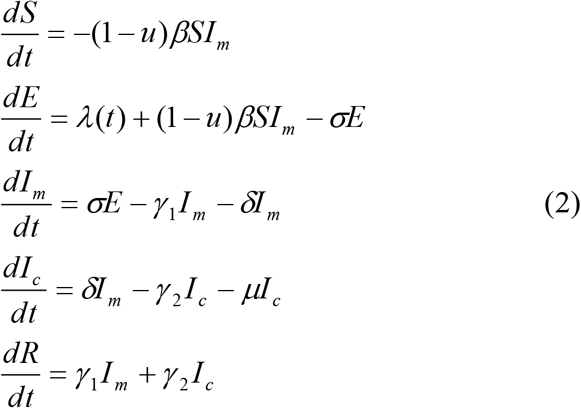

Following the same method described for system (1), the basic reproduction number for the system (2) can be obtained as 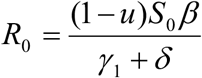 assuming the critically ill patients are fully isolated and biologically they are unable to transmit the virus anymore [17, 18].

#### Simulation of the *SEI*_*m*_*I*_*c*_*R* model (model 2) with critically ill patients

##### a) Sensitivity of the control measures

The aim of this model is to assess the efficacy of control measures, however, the timing of control measures is not included to the simulation. We vary the control parameter *u* which addresses the combined impact of government social distancing control measures [20]. The outcome of this simulation is given in Figure 4. It can be seen that as the efficacy of the control measure increases the epidemic curve is flatten and also it is possible to delay the peak of the outbreak so that the national health system is able to treat patients without getting overwhelmed [17].

**Figure 4:**
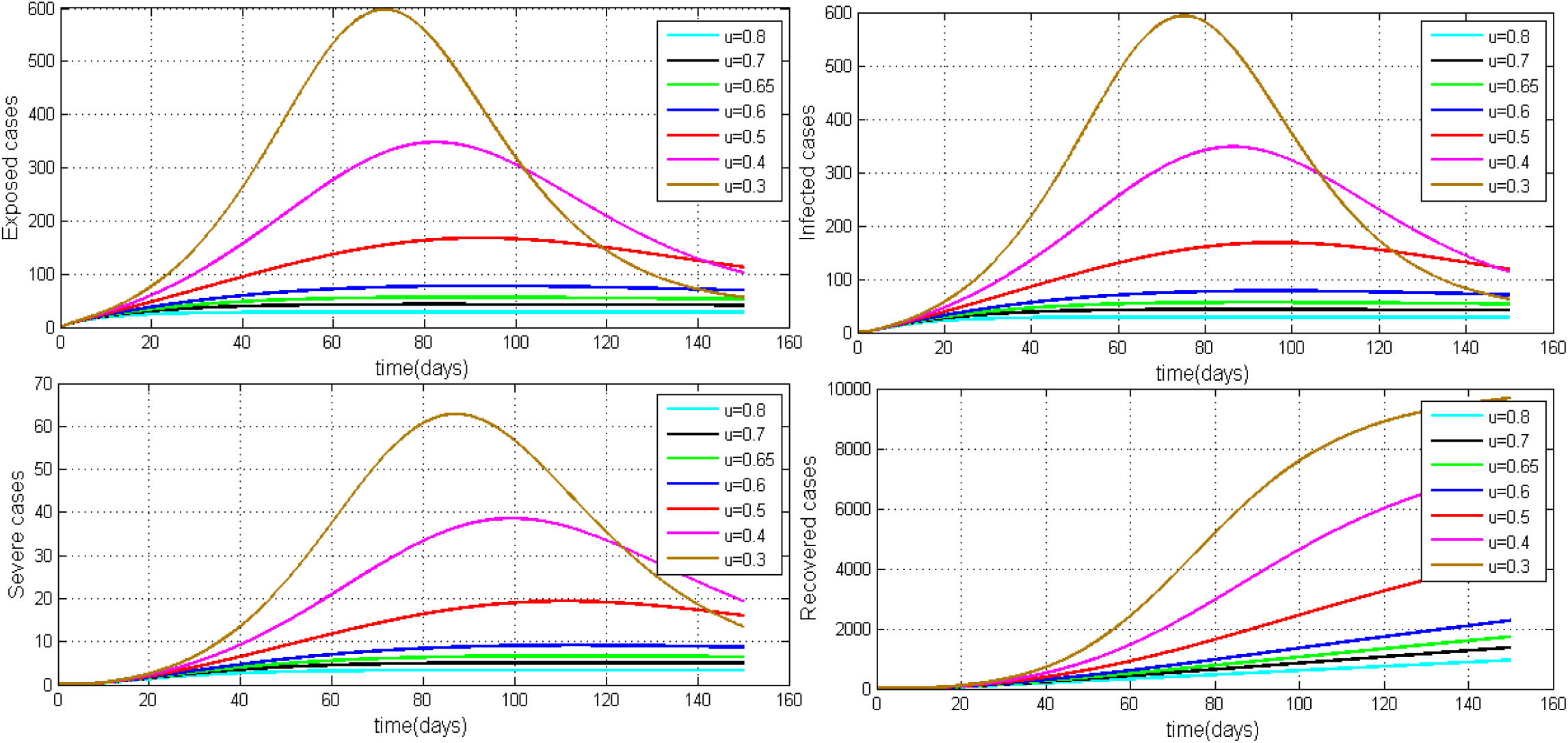
The simulation of *SEI*_*m*_*I*_*c*_*R* model considering the varying level of the control parameter *u*. The rest of the parameter values used for the simulation are *β* = 0.7, *γ*_1_ = 0.24, *γ*_2_ = 0.05, *μ* = 0.02, *σ* = 1/ 4, *δ* = 0.025/ 3 and *λ* = 0.000205 [13, 18].

Figure 5 shows how it is possible to reduce the maximum of the curve representing the patients who show mild symptoms and the maximum of the curve representing critically ill patients for the given period of time. The figure clearly shows that the size of this peak reduces as the combined control parameter *u* is increased from 30% to 80%.

**Figure 5:**
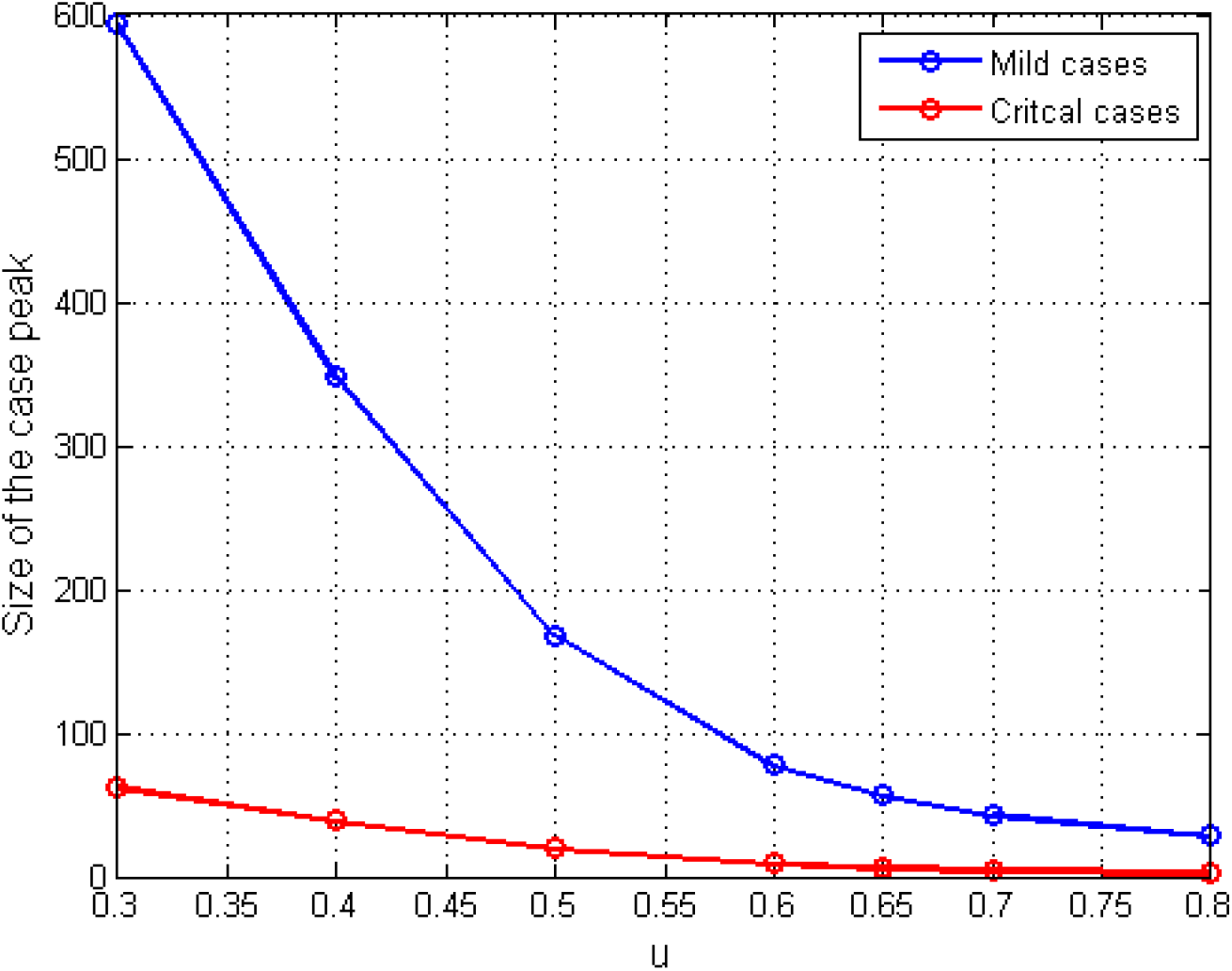
The change in the peak of mild cases and critical cases with respect to the combined control parameter *u*.

##### b) Sensitivity of the control of overseas exposed cases

The government of Sri Lanka decided to minimize the rate of imported exposed cases to the island five days after the first local COVID19 patient was identified. However, the authorities were flexible and allowed to arrive Sri Lankan citizens from selected countries for further about three days and thereafter, the decision was made to shut down the international airport for all arrivals to the country. We analyze the effect of this decision from the model simulation though the parameter *λ*. We define this parameter as a step function in time such that

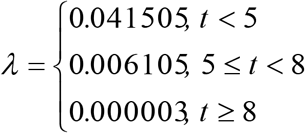

where *t* is the number of days after the first Sri Lankan case of COVID19 was identified by the health authorities in the country [12]. The outcome of this simulation is given in Figure 6.

**Figure 6:**
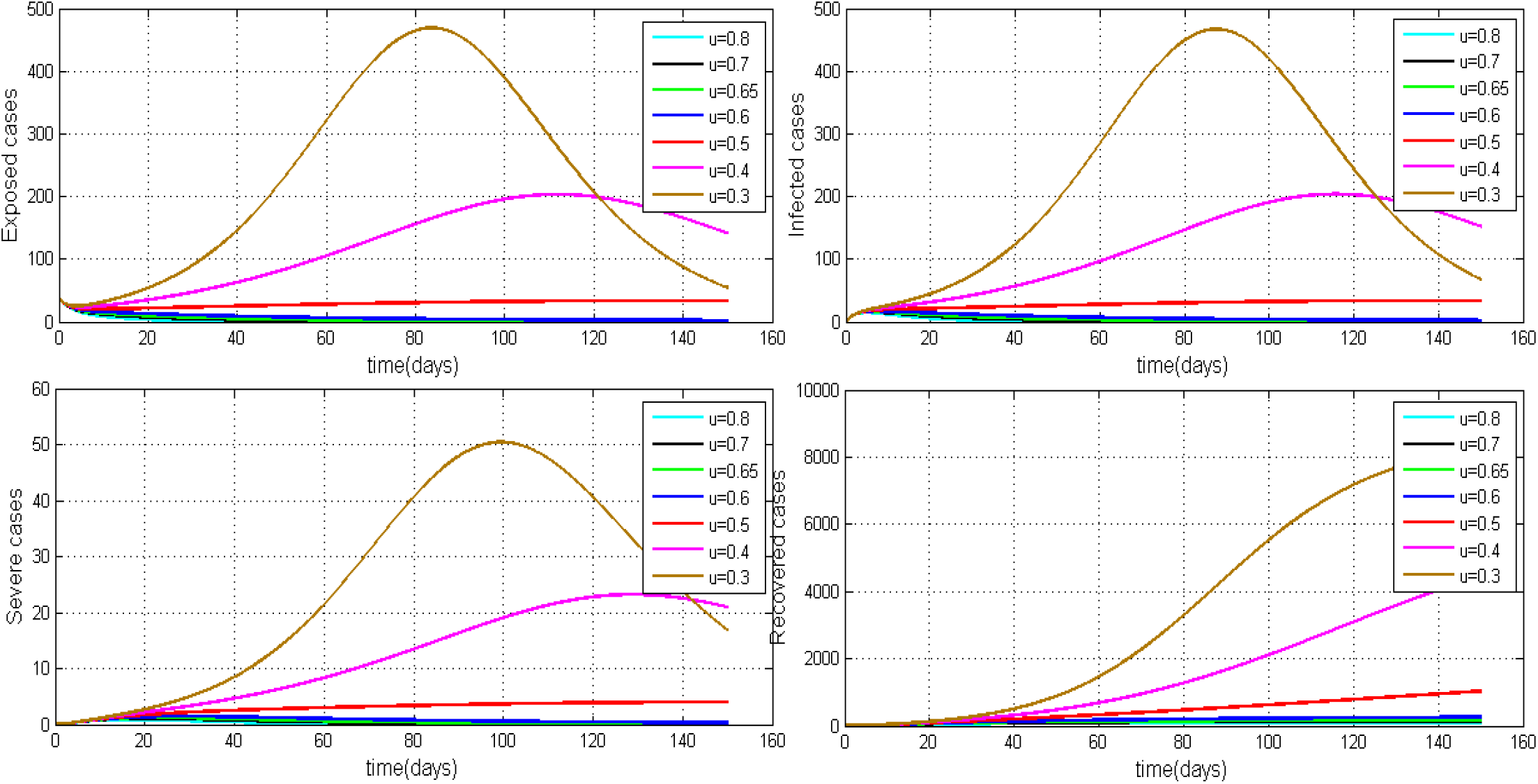
The simulation of *SEI*_*m*_*I*_*c*_*R* model considering the varying level of the control parameter *u* and the time effect of the decision to airport shut down. The rest of the parameter values used for the simulation are *β* = 0.7, *γ*_1_ = 0.24, *γ*_2_ = 0.05 *μ* = 0.02, *σ* = 1/ 4 and *δ* = 0.025/ 3 [13, 15, 17, 18].

Figure 7 shows how the peaks of the infected curved can be reduced with respect to the decision made to stop international arrivals who can be exposed to the virus.

**Figure 7:**
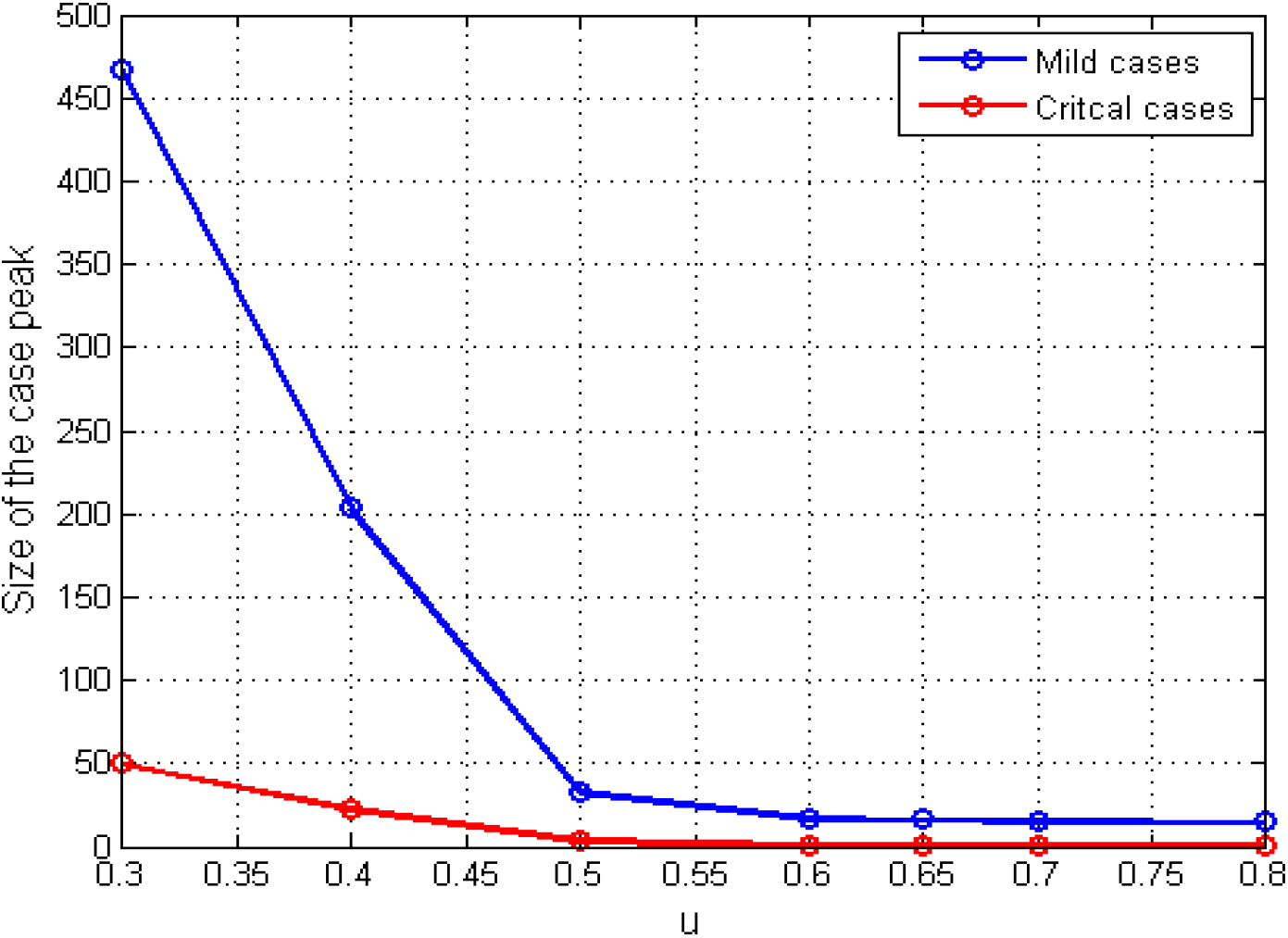
The change in the peak of mild cases and critical cases with respect to the combined control parameter *u* and the decision made to stop overseas exposed cases.

##### c) Sensitivity of the timing of implementing combined control measures

It is very critical to introduce social distancing control measures in correct points in time to minimize the burden of the COVID19 [15]. Most of the EU countries have found to have misjudged the scheduling of these measures and they have ended up with collapsing their health system causing significantly large number of COVID19 deaths. For this simulation, we vary the time in days from the date first local case was identified until the government decided to impose drastic social distancing control measures (80% efficacy). It is assumed that until this date, the authorities were very flexible and they are at mild level of restrictions (20% efficacy) [15, 17]. Figure 8 shows how the dynamic is changed with respective to this time parameter denoted by (Tr). Figure 9 demonstrates its sensitivity to both mild and critical cases peaks.

**Figure 8:**
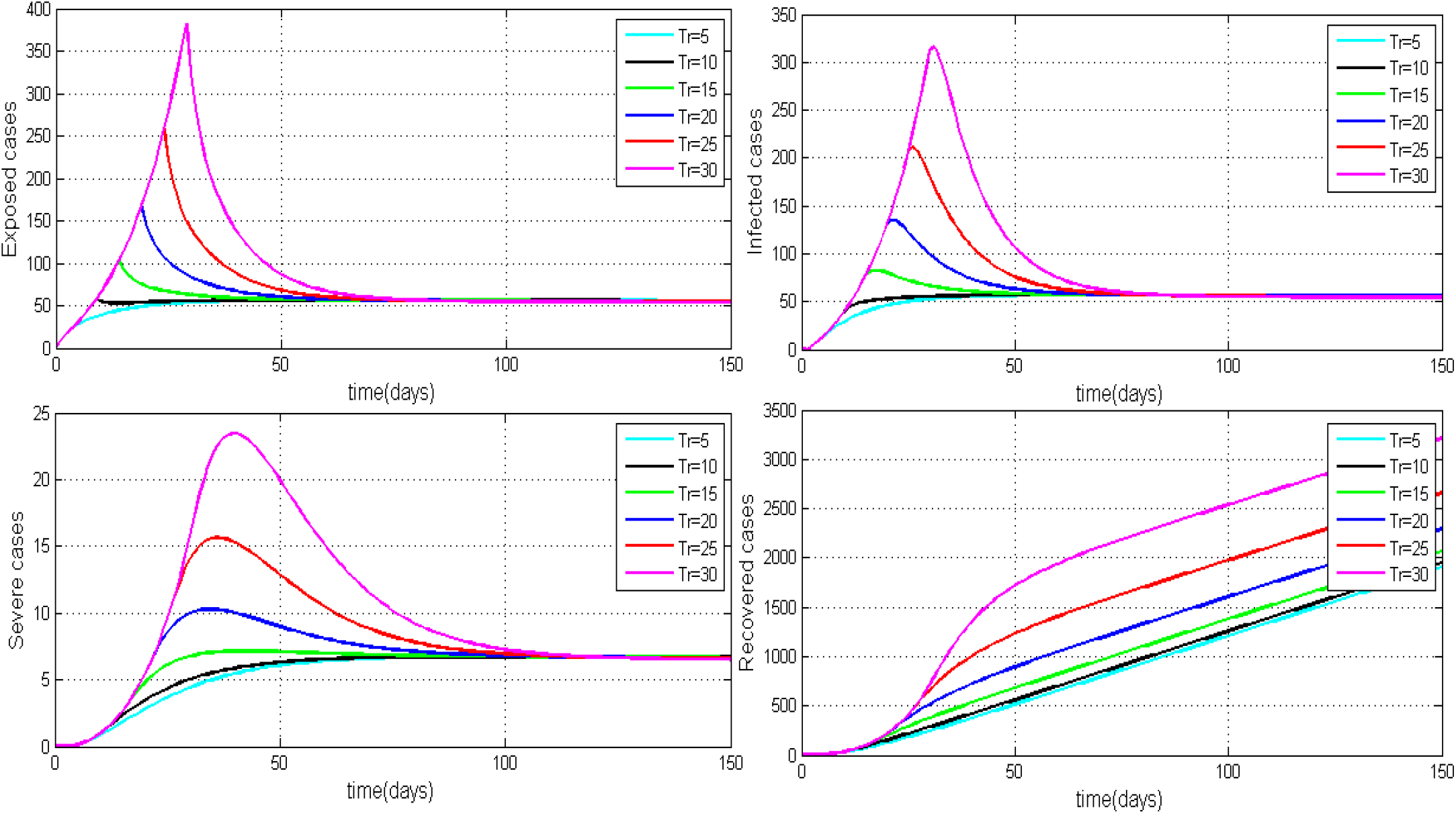
The simulation of *SEI*_*m*_*I*_*c*_*R* model considering the varying threshold time to impose strong social distancing control measures.

**Figure 9:**
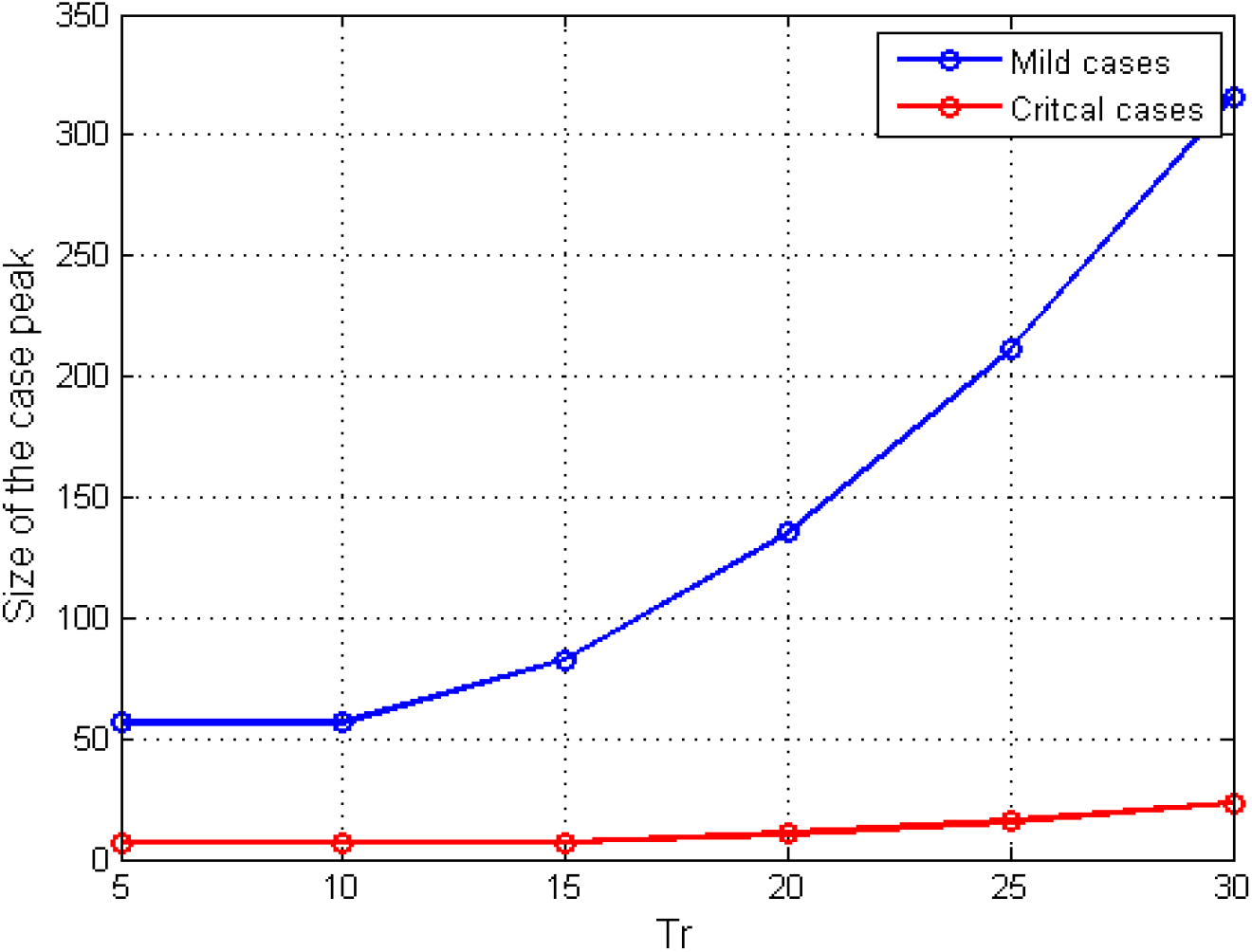
The change in the peak of mild cases and critical cases with respect to the threshold time to impose strong social distancing control measures.

According to Figure 8, it is very clear that this threshold value in time is very critical to minimize the burden to this highly contagious COVID19. It suggests that if the government had introduced 80% social distancing control measures within 5 days of the first local case was identified, we may have prevented the steeper growth of cases and reduce the size of the peak significantly. The model also demonstrates that if we wait nearly for 30 days to impose control measures then we are likely to experience a large peak of cases. Figure 9 illustrates the relationship between time to introduce measures and the peak of cases showing how rapidly it grows with each delay..

## Discussion and Conclusion

The simulations hypothetically show if policy makers toughen the control measures, they can delay the peak and thereby flatten the curve. This will enable the health care system in the country to cope with both mild and severely ill patients of the COVID19. However, these recommendations are with the proviso that the epidemic in the country is less than a month old and it’s too early to make firm medium-term predictions [19].

It is important and relevant to identify and evaluate the current efficacy level of average combine social distancing measures a percentage in Sri Lanka. The outbreak predication may be based on these values. With hindsight, one could argue that drastic control measures at the very beginning may have prevented the disease from getting established and spreading, e.g. banning all inward travel immediately after the first patient was detected. Of course, this was considered impractical with the available knowledge at the time. Similarly, severe restriction of movement of people with strict enforcement of curfews may eradicate the disease. However, practically people have to be allowed to buy essential provisions and curfews have to be lifted for a few houses. As a result, the efficacy level of restricted mobility is never going to be perfect and continuous with respect to time.

Considering the complexity and uncertainty, we have mathematically modelled the control parameter with respect to various social distancing factors. We have also predicted the outbreak with respect to identified determinants of various social distancing measures introduced in different ranges [20].

It is also vital to check the timing (schedule) of control measures in effect during the active outbreak period through the simulation as we have learnt that if the country passes a certain point then it has been unable to control the transmission regardless of how serious measures are being taken later. Finally, the literature from China suggests that found that only 20% of patients have developed the disease to critical stage requiring ICU care while others had less severe or mild or asymptomatic disease. Therefore, in the public health perspective, it would be helpful to come up with a way to model and predict the patients who may become critically ill and seeks ICU treatments considering multiple demographic factors.

## Data Availability

No data sets involved.

